# Alpha-Mannosidosis: Quantitative Oligosaccharide Biomarker Analysis in Dried Blood Spots for Diagnosis, Newborn Screening, and Treatment Monitoring

**DOI:** 10.64898/2026.09.16.26362975

**Authors:** Dominik Badekow, Michelle Jeschik, Benjamin Dreyer, Katharina Hoff, Anja Friederike Köhn, Mona Lindschau, Anna Baghdasaryan, Anibh Martin Das, Simone Harmeling, Xinying Hong, Pavel Ješina, Vassiliki Konstantopoulou, Francyne Kubaski, Alexander Laemmle, Florian Lagler, Thomas Lücke, Martin Magner, Lara Maleen Marten, Neslihan Önenli Mungan, Anke Schumann, Lina Verena Sevenich, Ronen Spiegel, Marcus Stange, Ali Tunç Tuncel, Sema Kalkan Uçar, Nicole Muschol, René Santer, Simona Murko

## Abstract

**Background:** Alpha-mannosidosis (AM) is a debilitating lysosomal storage disorder (LSD) caused by insufficient activity of alpha-mannosidase, an enzyme responsible for cleavage of alpha-linked mannose residues from N-glycan-type oligosaccharides during their degradation. As a result, specific oligosaccharides accumulate in tissues and are also detectable in body fluids. AM is a progressive multi-organ disease for which hematopoietic stem cell transplantation (HSCT) and enzyme replacement therapy (ERT) are therapeutic options, however, treatment results depend considerably on early diagnosis. Therefore, inclusion of AM into newborn screening panels would significantly contribute to improving therapeutic outcome.

**Methods:** A rapid and fully quantitative method for the specific AM oligosaccharide biomarker Man_2_GlcNAc in dried blood spots (DBS) using ultra-performance liquid chromatography coupled to tandem-mass spectrometry was developed. DBS from 420 controls, 27 AM patients (including five newborn screening samples), and 35 patients with other LSDs were analyzed.

**Results:** A distinct separation in biomarker concentrations was observed between unaffected individuals and untreated AM patients (0.1 – 0.8 *vs* 3.2 – 11.3 µmol Man_2_GlcNAc/L, *p*<0.0001). Furthermore, Man_2_GlcNAc concentrations significantly differed between untreated patients and those on ERT (*n* = 17, *p*<0.0001) or after HSCT (*n* = 15, *p*<0.0001), and between both treatment groups (*p*<0.0001). Moreover, we demonstrated that this method is appropriate for monitoring treatment efficacy by analysis of samples from individual patients before and after initiation of therapy. We retrospectively analyzed residual samples of dried blood spots (DBS) from routine newborn screening that had been stored from patients with later-confirmed AM, which resulted in clear separation from the concentrations observed in a newborn screening control cohort (7.7 – 11.3 *vs* 0.1 – 0.8 µmol Man_2_GlcNAc/L, *p*=0.0001).

**Conclusion:** We present a simple and rapid method for quantitative analysis of the specific AM oligosaccharide biomarker Man_2_GlcNAc, which is remarkably stable compared to enzymatic assays. Thus, the method is useful for selective screening in suspected cases as well as for monitoring the response to different forms of treatment. Furthermore, Man_2_GlcNAc has the potential to serve as a useful biomarker for newborn screening offering the prospect of earlier treatment initiation, improved therapeutic outcome and higher quality of life for AM patients.

## Background

Alpha-mannosidosis (AM, OMIM# 248500) is an inborn error of N-glycan degradation, caused by insufficient activity of lysosomal alpha-mannosidase (EC 3.2.1.24), an enzyme encoded by the *MAN2B1* gene in chromosomal region 19p13.13. Biallelic pathogenic variants within this gene result in this ultra-rare lysosomal storage disorder (LSD) with an estimated incidence of 1 in 500,000 to 1,000,000 [1,2,3]. Alpha-mannosidase is responsible for the cleavage of alpha-linked mannose residues from N-glycan-type oligosaccharides during lysosomal glycoprotein degradation [4].

Therefore, impaired alpha-mannosidase activity leads to the accumulation of specific oligosaccharides in tissues, causing lysosomal swelling and ultimately premature cell death and liberation of storage products into body fluids, including blood and urine [2,4]. Clinical manifestations of AM include dysmorphic features, skeletal abnormalities, organomegaly, and a variety of neurologic symptoms, such as intellectual disability, movement disorders, psychiatric manifestations and hearing impairment [1,2,3,4]. Without treatment, the prognosis is generally poor, with progressive disease limiting patients’ ability to live independently [3].

Treatments such as hematopoietic stem cell transplantation (HSCT) and enzyme replacement therapy (ERT) with velmanase alfa can mitigate organ manifestations and disease progression; however, treatment efficacy strongly depends on early initiation. Therefore, timely diagnosis is crucial to achieve optimal treatment outcome [5,6,7].

Current diagnostic approaches depend on the recognition of clinical features suggestive of AM. However, the variability of the disease often leads to delayed diagnosis. Biochemical diagnostics including assessment of nonspecific urinary oligosaccharides and/or alpha-mannosidase activity in leukocytes or dried blood spots (DBS) are the standard diagnostic tools. Often only if these analyses indicate the presence of AM is this verified by genetic testing to identify biallelic pathogenic variants in *MAN2B1* [8,9]. However, each biochemical analysis has at least one significant drawback. Frequently used non-quantitative methods, such as thin-layer chromatography, are not specific and not very sensitive [7,10]. While measuring enzyme activity in DBS is advantageous due to the ease of sampling and shipping, a principal problem is thermal stability. Although no data is available on enzyme stability in DBS, such problems have been reported with regard to alpha-mannosidase activity in cerebrospinal fluid samples [11]. Enzyme activity assessment from leukocytes is more reliable but considerably more complex and requires large amounts of blood to isolate a sufficient number of leukocytes [10].

To circumvent these shortcomings, a variety of new diagnostic methods based on liquid chromatography tandem-mass spectrometry (LC-MS/MS) detection have been proposed for AM and other LSDs. For AM, these methods are based on the analysis of specific oligosaccharide biomarkers, such as Man_2_GlcNAc (*i.e.,* Manα1→3Manβ1→4GlcNAc) and other mannosyl oligosaccharides with up to 9 mannose residues, which are all found to specifically accumulate in body fluids, such as blood and urine [4].

Although several methods of varying complexity have been proposed for screening for AM in urine, all are limited to providing a qualitative or semi-quantitative results [12,13,14,15,16,17,18,19,20,21]. Their non-quantitative nature is seen as a major drawback, particularly considering that the monitoring of biomarker concentrations in AM patients was identified as an important measure by an expert panel [15,22]. A fully quantitative method has only recently been proposed for Man_2_GlcNAc in serum and plasma combined with the semi-quantitative assessment of Man_3-6_GlcNAc [23].

Previous work by our group also addressed this problem by providing a fully quantitative method for the determination of Man_2_GlcNAc combined with the semi-quantitative assessment of Man_3-4_GlcNAc in spot urine samples [24]. By clearly distinguishing patients with AM from controls, both methods have emerged as promising tools for selective screening for AM and allow monitoring of treatment efficacy, with Man_2_GlcNAc being the most specific parameter.

Due to the simplicity of capillary blood sample collection and the special role of DBS in newborn screening, we additionally report on the successful development of an LC-MS/MS method for the quantitative detection of Man_2_GlcNAc in DBS. Further, we demonstrate that this method holds potential to be adapted for future application in newborn screening programs.

## Methods

### Samples

Blood samples for method development and validation were taken from a healthy volunteer (male, 26 - 30 years) and stored at -20 °C throughout the validation period, unless declared otherwise. Residual samples from six randomly selected, anonymized individuals from routine analyses were used to validate the matrix effect. DBS samples from 420 controls were randomly chosen from anonymized screening test cards from the Hamburg neonatal screening program for Northern Germany, provided that parents had consented to their use for research (NBS; *n* = 205, 100 males and 105 females), and residual samples from routine acylcarnitine analysis (*n* = 215, 119 males and 96 females) that were stored at room temperature for up to one month prior to biomarker analysis. The median age of the controls was 2 years with an interquartile range of 9 years. This control cohort was used for biomarker analysis in comparison to samples from AM patients of all age groups and for the generation of age-independent reference ranges. The NBS control cohort was used to compare NBS samples from AM patients and to generate preliminary reference ranges for NBS for AM. All samples from this cohort were collected between 36 and 72 hours after birth. According to § 12 of the Hamburg Hospital Act (HmbKHG), no separate consent is required for anonymized samples used for method development, validation, and the generation of reference ranges. DBS samples of AM patients and patients affected by other LSDs were stored at -80 °C. An overview of the AM patients included in this study, their age, sex, and treatment status as well as their molecular genetic findings and their residual α-mannosidase activity at the time of diagnosis, is provided in Table S1. A list of patients with other LSDs can be found in Table S2, including age, sex, and individual diagnosis (*see* Additional file 1).

We investigated DBS samples of unaffected controls, untreated AM patients (*n* = 14 samples from 13 individuals), AM patients on ERT (*n* = 17 samples from 12 individuals), AM patients after HSCT (*n* = 15 samples from 7 individuals), and patients with other LSDs (*n* = 36 samples from 35 individuals).

Among the samples of untreated AM patients, five were residuals from the neonatal screening testcard of the respective patients which we obtained after they had been diagnosed with AM. Hence, they had been stored at ambient temperature for 1 – 8 years in different newborn screening centers prior to inclusion in this study. Regarding the samples after HSCT, two of the patients (0079 and 0080) showed mixed chimerism and as a result, they were also on ERT at the time of sample collection. Their degree of hematological donor chimerism at the time of sampling is provided in Table S1. Likewise, two samples from patient 0082 were collected shortly after HSCT while still on ERT. All samples from patients who were still receiving ERT after HSCT at the time of sampling were classified into the HSCT cohort.

The diagnosis of all patients admitted to this study was confirmed by enzyme activity and/or genetic testing (= inclusion criteria for study samples).

### Materials

Acetonitrile (for UHPLC-MS) was purchased from Th. Geyer GmbH & Co. KG (Renningen, Germany). Water (HPLC Gradient grade) was obtained from Avantor Performance Materials, LLC (Center Valley, PA, USA). Formic acid (98 – 100 %) was supplied by Merck KGaA (Darmstadt, Germany). Man_2-4_GlcNAc (95 - 98 %) and the internal standard (IS) Man_2_GlcN-[1,2-^13^C_2_; 2-^2^H_3_]Ac (95 - 98 %) were obtained from Omicron Biochemicals, Inc. (South Bend, IN, USA).

### Sample analysis

DBS were processed with a manual punching device to generate a disc of 3.2 mm in diameter (= *punch*), which was transferred to a 1.5 mL microcentrifuge tube. Extraction solvent (100 µL), consisting of 10 % (*v*/*v*) formic acid in water and containing the IS (50 ng/mL), was added before the mixture was shaken thoroughly to fully submerge the punch and subsequently incubated for 10 min at room temperature. Thereafter, 400 µL of acetonitrile were added, resulting in a final IS concentration of 10 ng/mL in the sample extract, and the sample was vortexed for 5 seconds. Subsequently, sample extracts were centrifuged with a Mini Centrifuge C-1200 (Labnet International, Inc., Edison, NJ, USA) for 1 min and the supernatant was transferred into a glass vial and measured by ultra-performance LC (UPLC)-MS/MS using an external calibration calculated from the Man_2_GlcNAc signal response relative to the IS. LC-MS/MS parameters are provided in the Additional File 1. All study samples were analyzed in duplicate.

### Validation

The presented method was validated for linearity, lower limit of quantification (LLOQ), carry-over, limit of detection (LOD) and limit of quantification (LOQ), precision and accuracy, matrix effect, stability, and analytical specificity according to the ICH guideline M10 on bioanalytical method validation and study sample analysis [25].

Where necessary, deviations were made from the procedural recommendations and the acceptance criteria of the guideline to account for the lack of a blank matrix.

### Statistical analysis and visualization

Data acquisition was performed by MassLynx V4.2 SCN1024 in combination with TargetLynx XS V4.2 SCN1024 for peak integration and calculations. The quantitation was based on linear regression with a weighting factor of 1/x. Biomarker concentration was determined via external calibration based on isotope-labelled Man_2_GlcNAc as internal standard.

For data visualization and statistical analyses, we used Python 3 in a Jupyter Notebook 7.0.8 environment equipped with the packages pandas (version 2.2.3), matplotlib (version 3.10.0), seaborn (version 0.13.2), numpy (version 2.1.3), scipy (version 1.15.3), and statsmodels (version 0.14.4). Significance testing for differences between study cohorts was done by Whitney-Mann *U* test. Where necessary, we accounted for type I error due to multiple testing by Holm-Bonferroni adjustment of *p*-values.

## Results

### Method validation

A comprehensive outline of the validation process and results is provided in Additional file 1. While starting with three potential biomarkers (Man_2-4_GlcNAc), two of them (Man_3_GlcNAc and Man_4_GlcNAc) did not yield satisfactory validation results and were excluded from interpretation. For Man_2_GlcNAc, a linear relationship exists between the detector response and the analyte concentration in a range of 1 to 100 ng/mL (corresponding to 0.29 to 29 µmol/L Man_2_GlcNAc in the sample). This was demonstrated by triplicate measurement of an 11(+0)-point solvent calibration, resulting in an R² above 0.99 and randomly distributed residuals. Linearity in matrix was shown by assessment of a 6(+0)-point matrix calibration, which resulted in an R² of 0.986. The method is highly sensitive with an LOD for Man_2_GlcNAc below 0.1 µmol/L. The determination of accuracy and precision further highlights the good method performance for Man_2_GlcNAc, with accuracies ranging from 80 – 96 % and precisions below 14 % relative standard deviation. This finding is supported by a negligible matrix effect on quantification results. Notably, stability investigations demonstrate high stability of Man_2_GlcNAc at room temperature (20 °C) and under freezing conditions (-20 °C) for a period of at least 41 weeks.

### Man_2_GlcNAc concentrations in patients and controls

The individual biomarker concentrations of samples from AM patients and patients with other LSDs are provided in Tables S1 and S2, respectively (Additional file 1). Aside from the age that individual patients had reached, clinical information regarding the severity of the respective LSDs, including alpha-mannosidosis, was not available. An overview of the minimum and maximum concentrations of Man_2_GlcNAc observed in each cohort, including both NBS cohorts, is provided in Table 1.

**Table 1:** Biomarker concentration ranges in all study cohorts.

| Cohort | Sample size (n) | Range of Man<sub>2</sub>GlcNAc [μmol/L] |
| --- | --- | --- |
| Controls | 215 | 0.1 – 0.8 |
| AM Untreated | 13 | 3.2 – 11.3 |
| AM on ERT | 17 | 1.2 – 4.6 |
| AM after HSCT | 15 | 0.2 – 1.8* |
| Other LSDs | 36 | 0.1 – 0.7 |
| NBS Controls | 205 | 0.1 – 0.8 |
| NBS AM | 5 | 7.7 – 11.3 |

We determined the 99 % confidence reference intervals from the Man_2_GlcNAc concentrations of both control cohorts by interpolating the values at the 0.5^th^ and 99.5^th^ percentile, which results in a reference interval for unaffected individuals of 0.2 to 0.7 µmol/L for the general population and 0.2 to 0.8 µmol/L for newborns. No significant differences in biomarker concentration were observed based on sex (*data not shown*). Man_2_GlcNAc concentrations are marginally higher in samples from younger individuals. However, age-dependent differences within the control group are markedly smaller than those observed between controls and untreated AM patients. Hence, there is no benefit from age-adjusted reference intervals and we decided not to implement them.

The comparison of the Man_2_GlcNAc concentrations of the control cohort and the untreated AM patients results in a statistically highly significant difference (*p* < 0.0001; Figure 1). A comparison of patients with other LSDs and untreated AM patients also showed a statistically highly significant difference (*p* < 0.0001) (Figure 1) and it is worth noting that the range of biomarker concentration in patients with other LSDs is very similar to the range of the control cohort (Table 1). A statistically highly significant difference (*p* < 0.0001) was further observed for the concentrations of Man_2_GlcNAc of untreated AM patients and those who are on ERT (Figure 2). Man_2_GlcNAc concentrations are significantly higher in AM patients on ERT when compared to patients after HSCT (*p* < 0.0001; Figure 2). In addition, it is remarkable that the concentration of Man_2_GlcNAc in the samples from patients after HSCT, who did not exhibit mixed chimerism, is well within the range of the control cohort. The samples of the two patients with mixed chimerism and additional ERT on the other hand, show higher Man_2_GlcNAc concentrations than the reference interval and fall in the lower end or slightly below the concentration range of patients on ERT.

**Figure 1:**
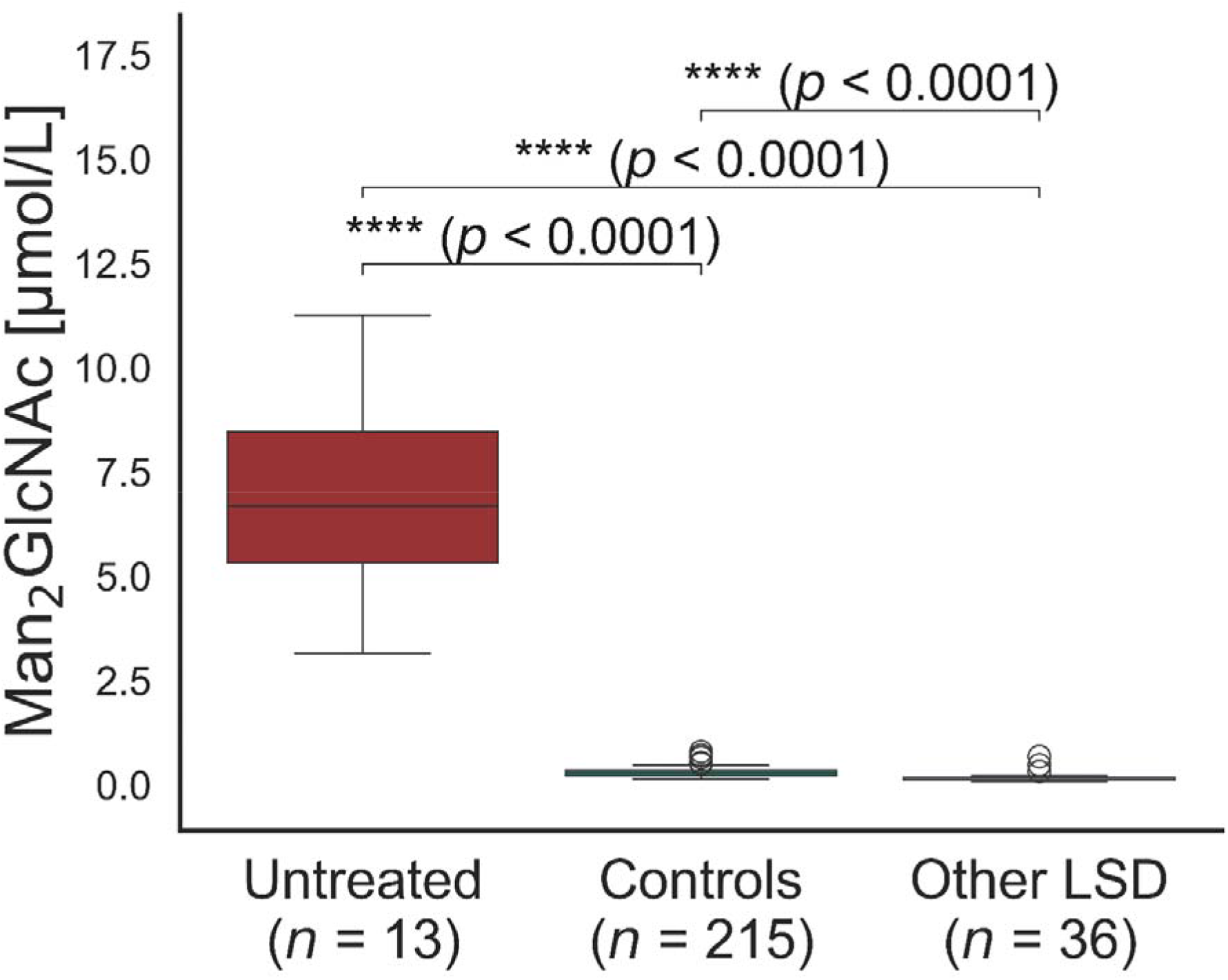
Diagnostic sensitivity and specificity. Box plots of the concentrations of Man2GlcNAc in samples of untreated AM patients, controls, and patients of other LSDs along with p-values of Whitney-Mann U tests after Holm-Bonferroni correction for type I error. AM, alpha-mannosidosis; ERT, enzyme replacement therapy; HSCT, hematopoietic stem cell transplantation; LSD, lysosomal storage disorder.

**Figure 2:**
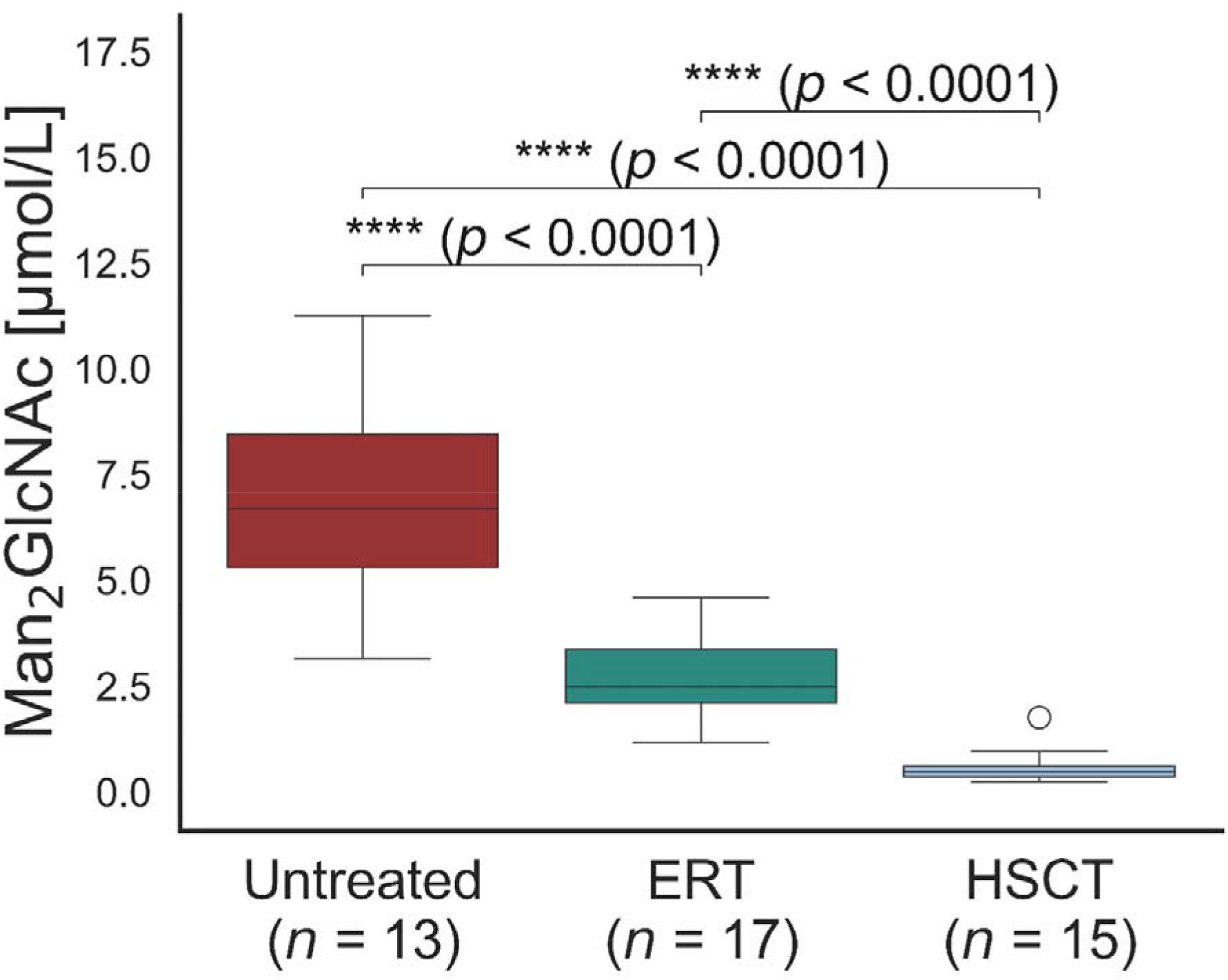
Biomarker concentration as a function of treatment type. Box plots of the concentrations of Man2GlcNAc in samples of untreated AM patients, patients receiving ERT, and patients after HSCT. p-values are the results of Whitney-Mann U tests after Holm-Bonferroni correction for type I error. AM, alpha-mannosidosis; ERT, enzyme replacement therapy; HSCT, hematopoietic stem cell transplantation.

### Man_2_GlcNAc concentration in neonatal DBS of AM patients

For this study, we were able to trace the neonatal testcard of five patients who were later diagnosed with AM. Residuals of these cards, which were usually kept at ambient temperature, were sent to our laboratory after different time periods (1 to 8 years).

Notably, the Man_2_GlcNAc concentrations of these neonatal samples were among the highest of all samples of untreated AM patients, resulting in a very strong separation from the NBS control cohort (> 9-fold; Table 1). We found a statistically highly significant difference (*p* = 0.0001) between the Man_2_GlcNAc concentrations of the samples from the NBS control cohort and NBS samples of AM patients (Figure 3).

**Figure 3:**
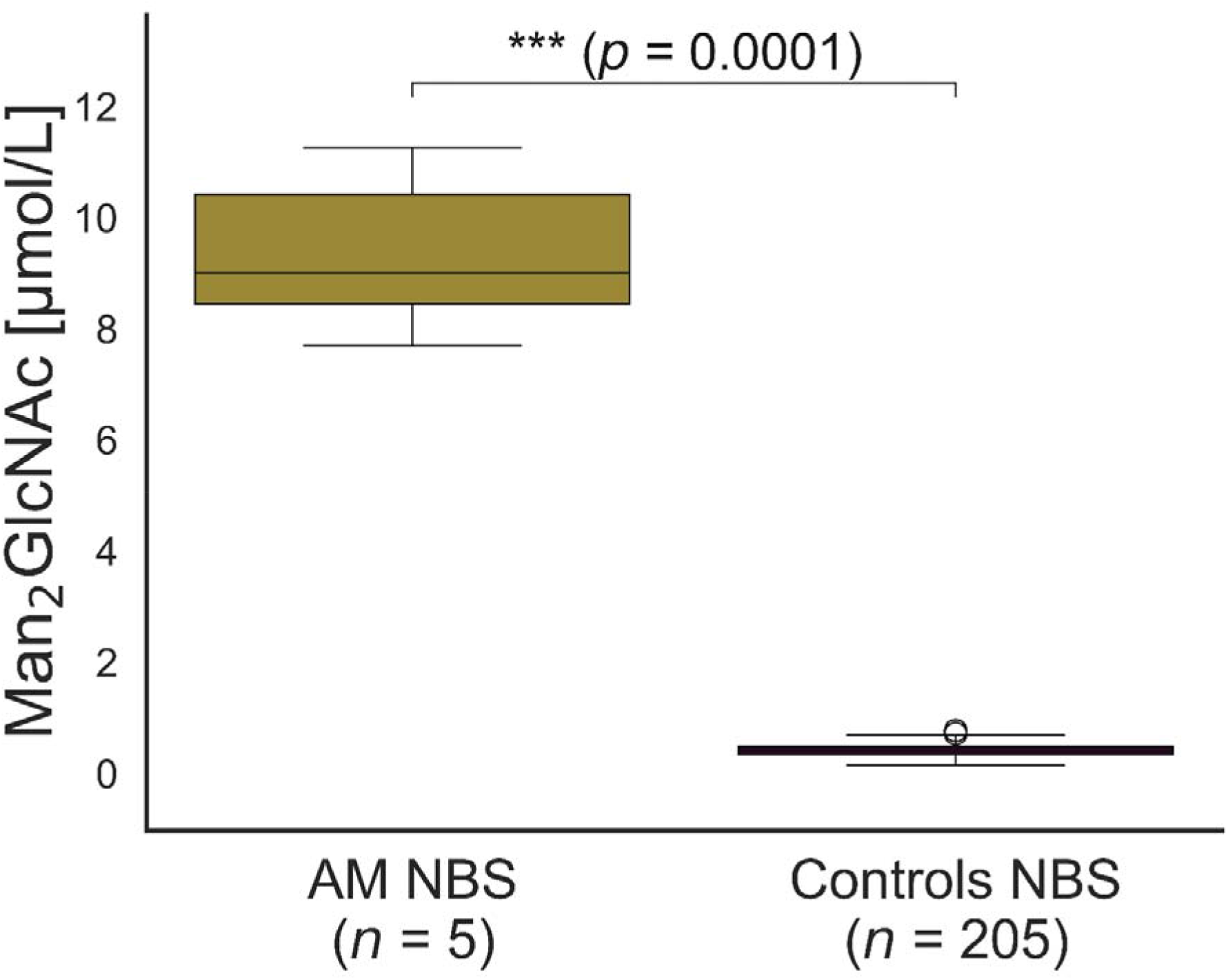
Biomarker concentration in neonatal samples.Box plots of the concentrations of Man2GlcNAc in newborn screening samples. AM, alpha-mannosidosis; NBS, newborn screening.

## Discussion

### Method validation

We successfully validated the newly developed UPLC-MS/MS method for the quantification of Man_2_GlcNAc in DBS for linearity and LLOQ, carry-over, LOD and LOQ, accuracy, precision, matrix effect, stability, and analytical specificity. In conclusion, this method is well-suited for the sensitive, accurate, and robust quantification of Man_2_GlcNAc in DBS. This oligosaccharide was previously considered an essential biomarker for AM, as it is not an intermediate product of normal catabolic metabolism of high-mannose, hybrid or complex N-linked oligosaccharides [4] – an assessment recently confirmed by our group as well as others [23,24].

### Biomarker concentrations for the diagnosis of alpha-mannosidosis

Evaluation of the reference range shows clear separation between the upper limit of the Man_2_GlcNAc reference range for controls and the lower limit of the concentrations observed in samples from untreated AM patients (> 3-fold). These results support the use of Man_2_GlcNAc as disease-specific biomarker for AM. On this basis and in accordance with the established reference intervals for unaffected individuals, we established a cut-off value for Man_2_GlcNAc in DBS of 1 µmol/L for the diagnosis of AM.

### Diagnostic sensitivity and specificity

As a result of the clear separation between the Man_2_GlcNAc concentrations in DBS of the control cohort and the cohort of untreated AM patients, all untreated AM patients were classified correctly by use of the afore-mentioned cut-off. Hence, this method’s diagnostic sensitivity is 100 %. Moreover, our assessment of diagnostic specificity also resulted in 100 %, since none of the 35 patients affected by other LSDs would have been classified as affected by AM. In conclusion, this method is appropriate for diagnosis of AM. In addition to the high diagnostic value of our method, the thermal stability of the biomarker allows samples to be transported over long distances without refrigeration, without causing a decrease in biomarker concentration and thus without compromising accuracy or adversely affecting the correct diagnosis.

### Treatment monitoring

The development of this method further pursued the objective to enable the monitoring of treatment efficacy by quantitative biomarker analysis. To assess this, we compared the Man_2_GlcNAc concentrations in the samples of untreated AM patients, patients on ERT, and patients after HSCT.

The ranges of untreated AM patients and those receiving ERT show some overlap (Figure 2). One could speculate that this is the result of different ages at the start of therapy and/or different treatment durations, however, our data analysis did not reveal any evidence to suggest this (*data not shown*). Overall, the Man_2_GlcNAc concentrations are generally lower in patients receiving ERT than in untreated patients, with some concentrations being as low as 1.2 µmol/L (*p* < 0.0001). The samples of patients after HSCT with full donor chimerism exhibited normalized levels of Man_2_GlcNAc concentrations (0.2 – 0.7 µmol/L). Therefore, the Man_2_GlcNAc concentrations of the patients after HSCT (with donor chimerism ≥ 90 %) are well-separated from the concentration range of samples from untreated AM patients and also from patients receiving ERT (*p* < 0.0001).

In few AM cases, we had the opportunity to analyze samples pre- and post-HSCT treatment (patients 0043 and 0082). The samples from patient 0043 were collected before, and up to 13 years after HSCT and allow for the assessment of long-term effects of HSCT on the biomarker concentration. Figure 4 shows a marked decrease of Man_2_GlcNAc in DBS after HSCT, which subsequently remains stable at a low concentration level. The samples from patient 0082 were taken before and few weeks after HSCT, when the patient was still receiving ERT during a transition period. The results show a very similar decrease of Man_2_GlcNAc as in patient 0043 (Figure 4), although it should be noted that the concurrent ERT may in part be responsible for this decrease. Nonetheless, the oligosaccharide biomarker concentrations are considerably lower than in patients treated exclusively with ERT. Hence, HSCT seems to lead to larger reductions of oligosaccharide biomarker concentration in DBS even within few weeks after transplantation. The results of the two patients who developed mixed chimerism with 30 and 47 % donor cells after HSCT showed higher Man_2_GlcNAc concentrations in DBS compared to those with full donor chimerism (Table S1), even though they were also receiving ERT at the time of sampling. This suggests that biomarker monitoring is sufficiently sensitive to detect changes in the course of therapy. This would mean that our results are consistent with findings of Kubaski et al. [23], who demonstrated that concentrations of Man_2_GlcNAc in plasma and serum decrease substantially following HSCT and can be used for treatment monitoring in patients with AM. However, additional studies are necessary – especially those that collect DBS data from more patients with more time-points to validate these findings. Lastly, Man_2_GlcNAc concentrations of patients receiving ERT (> 1.2 µmol/L) are higher than those in patients post-HSCT who did not experience mixed chimerism (0.2 – 0.7 µmol/L), demonstrating the ability of our method to sensitively detect differences between treatments. These differences may be explained by the inability of the replacement enzyme to cross the blood-brain barrier [26], leading to continued accumulation of Man_2_GlcNAc in brain tissue, which may subsequently be released into the bloodstream. In conclusion, our method clearly separates treated and untreated patients, it effectively helps with treatment monitoring, and it may also provide a hint on the degree of mixed chimerism after HSCT, even if patients receive ERT concurrently.

**Figure 4:**
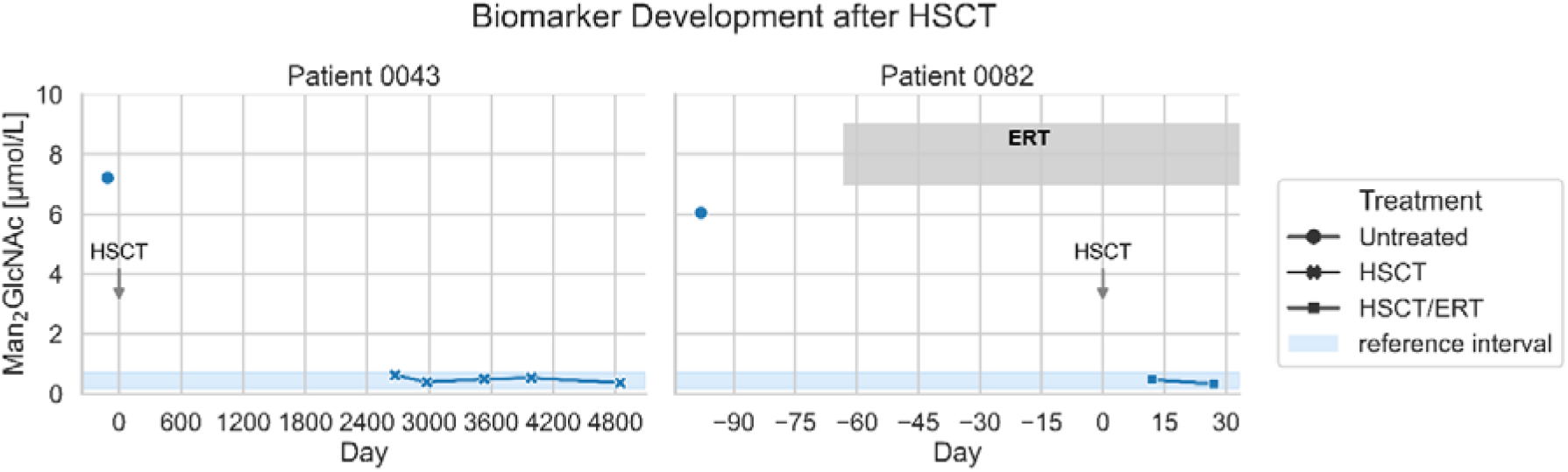
Impact of HSCT and HSCT with concurrent ERT on biomarker concentration. Development of the Man2GlcNAc concentration before and after HSCT in two patients – one with concurrent ERT and one without. Day 0 marks the date of HSCT. HSCT, hematopoietic stem cell transplantation; ERT, enzyme replacement therapy.

### Newborn screening

Currently, AM is not included in any official newborn screening panel. So far, it is merely a plan to add screening for AM to one pilot study with a flexible panel (ScreenPlus, New York) [27], where enzyme activity was chosen as the screening parameter (Melissa P. Wasserstein, *personal communication*). Nonetheless, experts regularly call for the inclusion of AM in newborn screening panels. Therefore, we aimed to investigate the potential of a biomarker-based newborn screening method for AM. To this end, we were able to identify five residual samples from newborn screening test cards of patients who were subsequently diagnosed with AM years after birth.

Unexpectedly, the separation achieved with this method between NBS samples from AM patients and a NBS control cohort was very good (> 9-fold; p = 0.0001; Table 1). We cannot ultimately rule out the possibility that these high Man_2_GlcNAc concentrations in DBS from newborns are the result of the degradation of higher molecular weight substances due to relatively long storage times under poorly controlled conditions. However, our own investigations into the stability of Man_2_GlcNAc in DBS over a period of almost one year did not reveal any increase over time which could support such a mechanism (Figure S2) and measured biomarker range in neonatal DBS is about in the range of the cohort of selectively diagnosed untreated patients.

Although it is generally accepted that AM patients do not show clinical symptoms at birth [28], accumulation of oligosaccharides in fetal and placental tissue could be demonstrated in a naturally occurring animal model [29]. Thus, it is to be expected that specific oligosaccharides also have entered the blood stream prenatally although no data is available for humans (*e.g*., from prenatal diagnostic approaches). Therefore, we were further interested in whether there is prenatal transport of accumulating oligosaccharides across the placenta from an affected fetus to the (heterozygous) mother. Oligosaccharides are hydrophilic substances that would require specific transport systems for transplacental transport. Maternal milk oligosaccharides have been detected in fetal blood; however, it is assumed that they are transported towards the fetus via active transport systems and not *vice versa* [30]. In summary, the nature of such transport mechanisms for oligosaccharides in the placenta have remained elusive and there is currently no scientific evidence that complex oligosaccharides other than milk oligosaccharides cross the human placenta in significant amounts. We therefore conclude that the detection of the AM-specific oligosaccharide Man_2_GlcNAc in neonatal DBS could be a valuable tool enabling early diagnosis of AM. Our method using a thermostable biomarker would certainly be advantageous over the enzymatic method currently used under study conditions. In other matrices, namely cerebrospinal fluid, alpha-mannosidase has proven to be relatively unstable [11].

The method presented could, in principle, be applied for newborn screening. In fact, because of the high linearity between biomarker concentration and detector response, a method based on the biomarker/IS-ratio for newborn screening should also be feasible.

To establish a method for NBS laboratories, however, further methodological adaptations are needed focusing on reducing run-time and increasing throughput potential, and the possibility of multiplexing with other biomarkers or enzymatic assays. Additionally, a larger number of NBS samples from both controls and affected patients needs to be analyzed to provide more reliable reference intervals.

Nonetheless, such a method would hold significant promise for AM patients, since early diagnosis is a key factor driving treatment efficacy [5,6,7].

### Limitations

Despite the important results that were obtained with the new biomarker detection method, our study has some limitations. One is the low overall number of available patient samples and especially the low number of samples per cohort, necessitating a combined analysis of cross-sectional and longitudinal data. This, however, is a result of the rarity of AM with only few known patients even in specialized centers. Moreover, the residual samples from newborn screening of AM patients have been stored for up to eight years – a time span for which no stability assessment has yet been performed.

## Conclusion

We present a rapid and simple LC-MS/MS method for selective screening and treatment monitoring of AM, which also proved effective as a first-tier for newborn screening of AM. The method is sensitive, robust, and accurate, and long-term stability of the AM biomarker Man_2_GlcNAc in DBS is notable.

The comparison of Man_2_GlcNAc concentrations in samples of untreated AM patients, healthy controls, and patients with other LSDs resulted in clear separations, highlighting diagnostic reliability. Additionally, we observed differences in biomarker concentrations between untreated AM patients, AM patients on ERT, and after HSCT, indicating the method’s suitability for monitoring treatment efficacy.

## List of abbreviations

AM: alpha-mannosidosis
DBS: dried blood spots
ERT: enzyme replacement therapy
HSCT: hematopoietic stem cell transplantation
IS: internal standard
LC: liquid chromatography
LLOQ: lower limit of quantification
LOD: limit of detection
LOQ: limit of quantification
LSD: lysosomal storage disorder
MS/MS: tandem-mass spectrometry
NBS: newborn screening

## Ethics approval and informed consent

Ethical approval for the study was granted by the ethics committee of the Medical Association Hamburg (Ärztekammer Hamburg, 06.02.2024, processing number 2023-101200-BO-ff). Informed consent was obtained from all study participants and, where applicable, their legal guardians prior to inclusion. All procedures were in accordance with the ethical standards of the responsible committees on human experimentation (institutional and national) and with the Helsinki Declaration of 1975, as revised in 2000.

## Consent for publication

Not applicable.

## Availability of data and materials

The datasets generated and analyzed during the current study are available in figshare repository, 10.6084/m9.figshare.32934854.

## Author’s contributions

DB conceptualized the research, developed the method and led the validation process, performed the experiments, analyzed and curated the data, wrote the python code for visualization of results, and wrote and edited the manuscript. MJ, BD, and KH supported method validation and edited the manuscript. AFK, ML, AB, AMD, SH, XH, PJ, VK, FK, AL, FL, TL, MM, LMM, NÖM, AS, LVS, RoS, MS, ATT, SKU, and NM provided essential resources, such as test samples and data from their patients, and edited the manuscript. NM additionally assisted in conceptualization and funding acquisition. ReS conceptualized the research, acquired funding, assisted in project administration and supervision, and wrote and edited the manuscript. SM (Guarantor) conceptualized the research, acquired funding, provided resources, was responsible for project administration and supervision, and edited the manuscript. All authors read and approved the final version of the manuscript and agree with submission. This work has not been published or submitted elsewhere.

## Additional Files

Additional file 1 (additional_file_1.docx): Additional information on study samples and method validation.

## Acknowledgements

The authors thank Oliver Blankenstein, Ipek Harman, Jakub Hodík, and Jeanette Klein for their efforts during sample acquisition.

