## Additional File 1 for "Alpha-Mannosidosis: Quantitative Oligosaccharide Biomarker Analysis in Dried Blood Spots for Diagnosis, Newborn Screening, and Treatment Monitoring"

**Alpha-Mannosidosis:**

### Samples

The following tables contain information on the patients included in this study. Table S1 lists all patients affected by AM, their treatment status, age at sampling, sex, and – if available – results of genetic analyses of the *MAN2B1* gene and residual enzymatic activity at time of diagnosis. For HSCT patients, degree of chimerism at the time of sampling is also provided, if available. For all AM patients of this study, biomarker concentrations, Man_2_GlcNAc measured in dried blood spots, are listed in the last column of this table. Table S2 lists all patients with other lysosomal storage disorders, including information on diagnosis, age at sampling, and sex. Also, the results of the quantification of Man_2_GlcNAc in DBS samples from these patients are listed here.

Patient and sample numbers are provided in the same format as they are used in our study cohort and as they appear in the sample lists and data files. The sample identifier is the combination of patient number and sample number in the format XXXX_XXXX. As an example, the first sample obtained from patient 2 in 2025 would be 0002_0125. It should be noted that some numbers are missing. These numbers refer to patients from whom we did not obtain DBS samples but other materials. Patient IDs are not known to anyone outside the research group.

**Table S1:** Overview of AM patients in this study including results of genetic analyses, residual enzymatic activity at time of diagnosis, and individual data for biomarker concentration (Man_2_GlcNAc) measured in dried blood spots.

|  | **Age range** | **Sex** | **Genotype *MAN2B1*** | | | | | | | | | | | | | | | | | | | | | | **Residual Activity *** | **Treatment Status** | **Chimerism** | **Biomarker Concentration** |
| --- | --- | --- | --- | --- | --- | --- | --- | --- | --- | --- | --- | --- | --- | --- | --- | --- | --- | --- | --- | --- | --- | --- | --- | --- | --- | --- | --- | --- |
| **Pt #** | [y] |  | ***First Allele***  ClinVar CADD GeneBe | | | | | | | | | | | ***Second Allele***  ClinVar CADD GeneBe | | | | | | | | | | | **[%]** |  |  | **Man_2_GlcNAc**  [µmol/L] |
| **0001** 0124 | 6-10 | f | c.212_215delCACA | | | | p.Thr71Metfs*85 | | | | | | | c.1830+1G>C | | | | | donor splice variant | | | | | | **1** | **ERT** | - | 2.5 |
| **0001** 0224 | 6-10 |  | **P** | | | **---** | | | | **P** | | | | **P** | **28** | | | | | | | **P** | | |  | **ERT** | - | 2.6 |
| **0006** 0124 | 11-15 | f | c.2151delG | | | | p.Pro718Argfs*48 | | | | | | | c.2151delG | | | | p.Pro718Argfs*48 | | | | | | | **0** | **ERT** | - | 1.2 |
|  |  |  | **---** | | | **---** | | | | **P** | | | | **---** | **---** | | | | | | | **P** | | |  |  |  |  |
| **0028** 0124 | 41-45 | m | c.2248C>T | | | | p.Arg750Trp | | | | | | | c.2183_2185delAGG | | | | p.Glu728del | | | | | | | **1** | **Untreated** | - | 6.7 |
|  |  |  | **P / LP** | | | **27** | | | | **P** | | | | **---** | **---** | | | | | | | **VUS** | | |  |  |  |  |
| **0035** 0124 | 31-35 | f | c.1830+1G>C | | | | donor splice variant | | | | | | | c.2248C>T | | | | p.Arg750Trp | | | | | | | **n/a** | **ERT** | - | 2.3 |
| **0035** 0125 | 31-35 |  | **P** | | | **28** | | | | **P** | | | | **P / LP** | **27** | | | | | | | **P** | | |  | **ERT** | - | 2.1 |
| **0038** 0117 | **0-5 ^†^** | m | c.2248C>T | | | | p.Arg750Trp | | | | | | | c.2248C>T | | | | p.Arg750Trp | | | | | | | **1** | **Untreated** | - | 8.5 |
| **0038** 0124 | 6-10 |  | **P / LP** | | | **27** | | | | **P** | | | | **P / LP** | **27** | | | | | | | **P** | | |  | **HSCT** | **100%** | 0.5 |
| **0039** 0124 | 36-40 | m | n/a | | | | | | | | | | | | | | | | | | | | | | **1** | **Untreated** | - | 4.1 |
|  |  |  | **---** | | | **---** | | | | **---** | | | | **---** | **---** | | | | | | | **---** | | |  |  |  |  |
| **0042** 0119 | 16-20 | m | n/a | | | | | | | | | | | | | | | | | | | | | | **2 / 9** ^•^ | **HSCT** | n/a | 0.7 |
| **0042** 0120 | 16-20 |  |  |  |  |  |  |  |  |  |  |  |  |  |  |  |  |  |  |  |  |  |  |  |  | **HSCT** | n/a | 0.4 |
| **0042** 0123 | 16-20 |  | **---** | **---** | | | | | | | **---** | | **---** | | | **---** | | | | | | | | **---** |  | **HSCT** | n/a | 0.6 |
| **0042** 0125 | 21-25 |  |  |  |  |  |  |  |  |  |  |  |  |  |  |  |  |  |  |  |  |  |  |  |  | **HSCT** | **99%** | 0.3 |
| **0043** 0112 | 0-5 | m | n/a | | | | | | | | | | | | | | | | | | | | | | **2 / 9** ^•^ | **Untreated** | n/a | 7.2 |
| **0043** 0119 | 6-10 |  |  |  |  |  |  |  |  |  |  |  |  |  |  |  |  |  |  |  |  |  |  |  |  | **HSCT** | n/a | 0.6 |
| **0043** 0120 | 6-10 |  |  |  |  |  |  |  |  |  |  |  |  |  |  |  |  |  |  |  |  |  |  |  |  | **HSCT** | n/a | 0.4 |
| **0043** 0122 | 11-15 |  | **---** | **---** | | | | | | | **---** | | **---** | | | **---** | | | | | | | **---** | |  | **HSCT** | n/a | 0.5 |
| **0043** 0123 | 11-15 |  |  |  |  |  |  |  |  |  |  |  |  |  |  |  |  |  |  |  |  |  |  |  |  | **HSCT** | n/a | 0.5 |
| **0043** 0125 | 11-15 |  |  |  |  |  |  |  |  |  |  |  |  |  |  |  |  |  |  |  |  |  |  |  |  | **HSCT** | **93%** | 0.4 |
| **0066** 0125 | 11-15 | m | c.2248C>T | | | | | p.Arg750Trp | | | | | c.259G>A | | | | | | | **p.Gly87Arg** | | | | | **0** |  | **-** | 3.2 |
|  |  |  | **P / LP** | **27** | | | | | | | **P / LP** | | **VUS** | | | **P** | | | | | | | **VUS** | |  |  |  |  |
| **0069** 0125 | 46-50 | f | c.2436+5G>A | | | | | donor splice variant | | | | | c.2887delG | | | | | | | p.Glu963Argfs*70 | | | | | **n/a** | **ERT** | **-** | 1.8 |
|  |  |  | **P / LP** | **35** | | | | | | | **LP** | | **VUS** | | | **---** | | | | | | | **LP** | |  |  |  |  |
| **0072** 0225 | 41-45 | m | c.2670_2671delAG | | | | | | p.Gly891Alafs*39 | | | | c.2670_2671delAG | | | | | | | | p.Gly891Alafs*39 | | | | **n/a** | **ERT** | - | 3.9 |
| **0072** 0325 | 41-45 |  | **P** | | **---** | | | | | | | **P** | **P** | | | | **---** | | | | | | | **P** |  | **ERT** | - | 2.7 |
| **0072** 0425 | 41-45 |  |  |  |  |  |  |  |  |  |  |  |  |  |  |  |  |  |  |  |  |  |  |  |  | **ERT** | - | 2.5 |
| **0073** 0125 | 0-5 | m | c.1694delT | | | | | | p.Leu565Argfs*16 | | | | c.1694delT | | | | | | | | p.Leu565Argfs*16 | | | | **0** | **ERT** | - | 3.4 |
| **0073** 0225 |  |  | **P** | | **---** | | | | | | | **P** | **P** | | | | **---** | | | | | | | **P** |  | **ERT** | - | 4.0 |
| **0074** 0121 | **0-5 ^†^** | m | c.418C>T | | | | | p.Arg140* | | | | | c.1349G>T | | | | | | | p.Ser450Ile | | | | | **0-14 /0/0** ^••^ | **Untreated** | - | 11.3 |
| **0074** 0123 | 0-5 |  | **P / LP** | | **40** | | | | | | | **P** | **VUS** | | | | **15** | | | | | | | **VUS** |  |  |  |  |
| **0075** 0125 | 11-15 | m | c. 856G>A | | | | | p. Glu286Lys | | | | | c. 856G>A | | | | | | | p. Glu286Lys | | | | | **0** | **ERT** | - | 2.3 |
|  |  |  | **LP** | | **23** | | | | | | | **LP** | **LP** | | | | **23** | | | | | | | **LP** |  |  |  |  |
| **0076** 0125 | 16-20 | m | c. 856G>A | | | | | p. Glu286Lys | | | | | c. 856G>A | | | | | | | p. Glu286Lys | | | | | **0** | **Untreated** | - | 4.3 |
|  |  |  | **LP** | | **23** | | | | | | | **LP** | **LP** | | | | **23** | | | | | | | **LP** |  |  |  |  |
| **0077** 0125 | 0-5 | f | c. 308C>T;  c.496G>T | | | | | p. Ser103Leu;  p. Gly166Cys | | | | | c. 308C>T;  c.496G>T | | | | | | | p. Ser103Leu;  p. Gly166Cys | | | | | **6.2** | **ERT** | - | 4.6 |
|  |  |  | **P_ VUS; ---** | | **33; 27** | | | | | | | **VUS; VUS** | **P_ VUS; ---** | | | | **33; 27** | | | | | | | **VUS; VUS** |  |  |  |  |
| **0078** 0125 | 6-10 | f | c. 308C>T;  c.496G>T | | | | | p. Ser103Leu;  p. Gly166Cys | | | | | c. 308C>T;  c.496G>T | | | | | | | p. Ser103Leu;  p. Gly166Cys | | | | | **0** | **ERT** | - | 3.9 |
|  |  |  | **P_ VUS; ---** | | **33; 27** | | | | | | | **VUS; VUS** | **P_ VUS; ---** | | | | **33; 27** | | | | | | | **VUS; VUS** |  |  |  |  |
| **0079** 0125 | 11-15 | m | c.1358 C>A | | | | | p.Ser453Tyr | | | | | c.1358 C>A | | | | | | | p.Ser453Tyr | | | | | **11** | **HSCT ‡** | **47%** | 1.0 |
|  |  |  | **VUS** | | **24** | | | | | | | **LP** | **VUS** | | | | **24** | | | | | | | **LP** |  |  |  |  |
| **0080** 0125 | 6-10 | f | c.1394_1395delTT | | | | | p.Leu465Argfs*51 | | | | | c.1321delG | | | | | | | p.Ala441Leufs*36 | | | | | **5** | **HSCT ‡** | **30%** | 1.8 |
|  |  |  | **---** | | **---** | | | | | | | **P** | **---** | | | | **---** | | | | | | | **P** |  |  |  |  |
| **0081** 0125 | 0-5 | f | c.631-1G>C | | | | | acceptor splice variant | | | | | c.631-1G>C | | | | | | | acceptor splice variant | | | | | **2** | **Untreated** | - | 5.3 |
|  |  |  | **---** | | **35** | | | | | | | **P** | **---** | | | | **35** | | | | | | | **P** |  |  |  |  |
| **0082** 0124 | **0-5 ^†^** | m | c.2248C>T | | | | | p.Arg750Trp | | | | | c.2398G>T | | | | | | | p.Gly800Trp | | | | | **1,4** | **Untreated** | n/a | 9.0 |
| **0082** 0225 | 0-5 |  |  |  |  |  |  |  |  |  |  |  |  |  |  |  |  |  |  |  |  |  |  |  |  | **Untreated** | n/a | 6.0 |
| **0082** 0725 | 0-5 |  | **P / LP** | **27** | | | | | | | **P** | | **LP** | | | **26** | | | | | | | | **P** |  | **HSCT ‡** | **100%** | 0.5 |
| **0082** 0825 | 0-5 |  |  |  |  |  |  |  |  |  |  |  |  |  |  |  |  |  |  |  |  |  |  |  |  | **HSCT ‡** | **100%** | 0.3 |
| **0083** 0122 | **0-5 ^†^** | f | c.1126G>A | | | | | p.Asp376Asn | | | | | c.1970C>T | | | | | | | p.Ser657Leu | | | | | **0.3** | **Untreated** | - | 10.4 |
|  |  |  | **VUS** | | **33** | | | | | | | **VUS** | **VUS** | | | | **26** | | | | | | | **VUS** |  |  |  |  |
| **0086** 0121 | **0-5 ^†^** | f | c.2248C>T | | | | | p.Arg750Trp | | | | | c.1313_1332dup | | | | | | | p.His445Argfs*39 | | | | | **0.3** | **Untreated** | - | 7.7 |
| **0086** 0125 | 0-5 |  | **P / LP** | | **27** | | | | | | | **P** | **---** | | | | **---** | | | | | | | **P** |  | **HSCT** | n/a | 0.2 (<LOQ) |
| **0090** 0126 | 11-15 | m | c.1183C>T | | | | | p.Arg395Trp | | | | | c.2248C>T | | | | | | | p.Arg750Trp | | | | | **2.2** | **ERT** | - | 2.1 |
|  |  |  | **---** | | **31** | | | | | | | **VUS** | **P / LP** | | | | **27** | | | | | | | **P** |  |  |  |  |
| **0091** 0126 | 16-20 | m | c.293dupA | | | | | p.Tyr99Valfs*62 | | | | | c.233T>C | | | | | | | p.Leu78Pro | | | | | **1.4** | **ERT** | - | 1.4 |
|  |  |  | **P** | | **---** | | | | | | | **P** | **VUS** | | | | **31** | | | | | | | **LP** |  |  |  |  |
| **0092** 0126 | 16-20 | m | c.293dupA | | | | | p.Tyr99Valfs*62 | | | | | c.293dupA | | | | | | | p.Tyr99Valfs*62 | | | | | **0.8** | **ERT** | - | 1.8 |
|  |  |  | **P** | | **---** | | | | | | | **P** | **P** | | | | **---** | | | | | | | **P** |  |  |  |  |

*AM*, alpha-mannosidosis; *ERT,* enzyme replacement therapy; *HSCT,* hematopoietic stem cell transplantation.

In addition to the genotype the variant classification according to ClinVar, the CADD score, and the summary assessment in accordance with GeneBe are provided with ‘VUS’, ‘LP’ and ‘P’ representing variants of unknown significance, likely pathogenic and pathogenic variants, respectively.

*n/a*, not available

**^†^**, residual newborn screening sample

*, from different materials

^•^, in dried blood spots and leukocytes, respectively

^••^, in dried blood spots, leukocytes and plasma, respectively

**^‡^**, patient received concurrent ERT at time of sampling

##### **Table S2:** Overview of patients with other lysosomal storage diseases in this study.

| **Patient number** | **Sample number** | **Lysosomal storage disease** | **Age range**  [years] | **Sex** | **Biomarker Concentration**  Man_2_GlcNAc  [µmol/L] |
| --- | --- | --- | --- | --- | --- |
| 0002 | 0124 | MPS I | 0-5 | male | 0.1 (<LOQ) |
| 0003 | 0124 | Gaucher disease | 0-5 | male | 0.1 (<LOQ) |
| 0004 | 0124 | Fabry disease | 51-55 | male | 0.1 (<LOQ) |
| 0005 | 0124 | Pompe disease | 11-15 | female | 0.1 (<LOQ) |
| 0007 | 0124 | MPS VI | 0-5 | male | 0.1 (<LOQ) |
| 0009 | 0124 | Pompe disease | 36-40 | female | 0.7 |
| 0010 | 0124 | Fabry disease | 11-15 | female | 0.2 (<LOQ) |
| 0011 | 0124 | Fabry disease | 66-70 | female | 0.1 (<LOQ) |
| 0012 | 0124 | Pompe disease | 51-55 | female | 0.1 (<LOQ) |
| 0013 | 0124 | Fabry disease | 26-30 | male | 0.2 (<LOQ) |
| 0014 | 0124 | Fabry disease | 41-45 | female | 0.2 (<LOQ) |
| 0015 | 0124 | MPS IIIA | 11-15 | male | 0.2 (<LOQ) |
| 0016 | 0124 | MPS IIIA | 16-20 | male | 0.2 (<LOQ) |
| 0017 | 0124 | MPS I | 6-10 | male | 0.2 (<LOQ) |
| 0019 | 0124 | Fabry disease | 46-50 | male | 0.1 (<LOQ) |
| 0020 | 0124 | Fabry disease | 11-15 | male | 0.1 (<LOQ) |
| 0021 | 0124 | Gaucher disease | 16-20 | female | 0.2 (<LOQ) |
| 0023 | 0124 | MPS I | 26-30 | female | 0.1 (<LOQ) |
| 0024 | 0124 | MPS I | 31-35 | female | 0.1 (<LOQ) |
| 0025 | 0124 | Schindler disease | 6-10 | male | 0.1 (<LOQ) |
| 0025 | 0224 | Schindler disease | 6-10 | male | 0.1 (<LOQ) |
| 0026 | 0124 | Pompe disease | 35-40 | female | 0.2 (<LOQ) |
| 0027 | 0124 | Gaucher disease | 31-35 | female | 0.1 (<LOQ) |
| 0029 | 0124 | MPS VI | 51-55 | female | 0.2 (<LOQ) |
| 0030 | 0124 | MPS II | 11-15 | male | 0.1 (<LOQ) |
| 0031 | 0124 | GM2 gangliosidosis | 0-5 | female | 0.1 (<LOQ) |
| 0033 | 0124 | Pompe disease | 6-10 | female | 0.1 (<LOQ) |
| 0051 | 0125 | ClC-7 hyperactivity ^∞^ | 6-10 | male | 0.2 (<LOQ) |
| 0052 | 0125 | Mucolipidosis II | 6-10 | female | 0.3 |
| 0053 | 0125 | Fucosidosis | 6-10 | male | 0.1 (<LOQ) |
| 0058 | 0125 | MPS II | 16-20 | male | 0.1 (<LOQ) |
| 0060 | 0125 | MPS VI | 56-60 | female | 0.5 |
| 0063 | 0125 | Fabry disease | 71-75 | male | 0.1 (<LOQ) |
| 0065 | 0125 | MPS I | 0-5 | male | 0.1 (<LOQ) |
| 0070 | 0125 | MPS IVB | 16-20 | male | 0.2 (<LOQ) |
| 0071 | 0125 | MPS IVB | 26-30 | male | 0.2 (<LOQ) |

*MPS,* mucopolysaccharidosis; *ClC-7,* chloride co-transporter-7.

^∞^ patient 1 in reference [1].

### Sample analysis

In the following section, the method parameters for LC-MS/MS analysis of the three oligosaccharide biomarkers Man_2_GlcNAc, Man_3_GlcNAc, and Man_4_GlcNAc are reported. As a result of method validation, it turned out that Man_3_GlcNAc and Man_4_GlcNAc are not reliably quantified by this method (*see* Method validation) and we made a conscious decision not to pursue this objective further. This decision was based on the increased workload that would have resulted from more extensive sample preparation as well as higher costs that would have been incurred by the need for specifically synthesized isotope-labelled standards for Man_3&4_GlcNAc. Notably, in previous studies quantifying the three oligosaccharide biomarkers in blood or urine, Man_2_GlcNAc has been the most abundant and the most reliable when it comes to making a diagnosis [2,3].

#### Ultra-performance liquid chromatography tandem-mass spectrometry

The quantitative analysis was conducted on a Waters ACQUITY UPLC^®^ I-Class coupled to a Waters Xevo TQ-S Micro triple quadrupole mass spectrometer. The analytes were separated on an ACQUITY UPLC^®^ Glycan BEH Amide column (130 Å, 1.7 µm, 2.1 mm × 150 mm, Waters, Eschborn, Germany) in combination with an ACQUITY UPLC^®^ Glycan BEH Amide pre-column (130 Å, 1.7 µm, 2.1 mm × 5 mm, Waters, Eschborn, Germany). Column temperature was set to 40 ± 5.0 °C. The mobile phase consisted of acetonitrile (eluent A) and water (eluent B), to both of which 0.1 % (*v/v*) formic acid was added. A linear gradient elution according to Table S3 was performed over a total run time of six minutes.

##### **Table S3:** Elution gradient settings.

| Run time  [min] | Eluent A  [%] | Eluent B  [%] | Flow rate  [mL/min] |
| --- | --- | --- | --- |
| 0 | 80 | 20 | 0.4 |
| 1 | 80 | 20 |  |
| 1.1 | 62 | 38 |  |
| 3.1 | 52 | 48 |  |
| 4.5 | 30 | 70 |  |
| 5 | 80 | 20 |  |
| 6 | 80 | 20 |  |

Injection volume was set to 10 µL. The MS/MS parameters are provided in Table S4. To reduce the strain of the matrix components on the MS/MS detector, dynamic multiple reaction monitoring was employed.

##### **Table S4**: MS/MS parameters for the detection of Man_2-4_GlcNAc.

| Analyte | Precursur Ion  [m/z] | Product Ion [m/z] | Dwell Time  [s] | Cone Voltage [V] | Collision Energy  [V] |
| --- | --- | --- | --- | --- | --- |
| Man_2_GlcNAc | 546.2 | 204.0 | 0.065 | 35 | 18 |
|  |  | 222.0 |  |  | 9 |
| Man_2_GlcNAc-IS | 551.2 | 209.0 |  | 35 | 18 |
|  |  | 227.0 |  |  | 9 |
| Man_3_GlcNAc | 708.3 | 204.0 |  | 40 | 25 |
|  |  | 222.0 |  |  | 14 |
| Man_4_GlcNAc | 870.3 | 204.0 |  | 70 | 30 |
|  |  | 222.0 |  |  | 18 |

*MS/MS,* tandem-mass spectrometry; *m/z,* mass-to-charge ratio;

*Man_2_GlcNAc-IS,* Man_2_GlcN-[1,2-^13^C_2_; 2-^2^H_3_]-Ac.

### Method validation

Method validation was conducted according to the ICH guideline M10 on bioanalytical method validation and study sample analysis supplied by the Committee for Medicinal Products for Human Use, European Medical Agency [4], with modifications where necessary. The method was validated for linearity and lower limit of quantification (LLOQ), carry-over, limit of detection (LOD) and quantification (LOQ), precision and accuracy, matrix effect, stability, and analytical specificity.

During method development, it was found that all DBS samples contain small amounts of the analytes. Therefore, no blank samples were available and instead, the sample of a healthy volunteer (male, 29) was used for method validation. The following sections give a detailed overview of the validation process and its results.

#### Linearity and lowest limit of quantification

Since no blank matrix is available for the three biomarkers, linearity was assessed in solvent for each of the three oligosaccharide biomarkers (Man_2_GlcNAc, Man_3_GlcNAc, and Man_4_GlcNAc) by independent and randomized triplicate analysis of an 11(+0)-point calibration curve ranging from 1 – 100 ng/mL. The concentration of the IS was kept constant at 10 ng/mL. Linear regression was performed, and linearity was assessed by R² (acceptance criterion: > 0.99), as well as visual evaluation of linearity and random distribution of residuals. Additionally, the linearity of the relationship between biomarker concentration and detector response from the analytes was assessed in matrix by adding seven different biomarker concentrations (0 ng/mL, 2 ng/mL, 20 ng/mL, 40 ng/mL, 60 ng/mL, 80 ng/mL, and 100 ng/mL) to an EDTA-blood sample of the healthy volunteer, which was subsequently applied to the newborn screening filter paper. After sample measurement, linear regression was performed and assessed by R² (acceptance criterion: > 0.98).

The results are shown in Figure S1. Linearity in solvent was confirmed in the range of 1 to 100 ng/mL for all three biomarkers. The R² is above 0.99 in solvent and above 0.98 in matrix, meeting both acceptance criteria (Table S5). The residuals of all three analytes are randomly distributed. Thus, a linear relationship between the detector response and the analyte concentration has been confirmed for all three analytes in solvent and matrix.

The LLOQ is calculated as the biomarker concentration in sample equal to the lowest calibration level. Therefore, the LLOQ is 0.29 µmol/L, 0.22 µmol/L, and 0.18 µmol/L for Man_2_GlcNAc, Man_3_GlcNAc, and Man_4_GlcNAc, respectively.

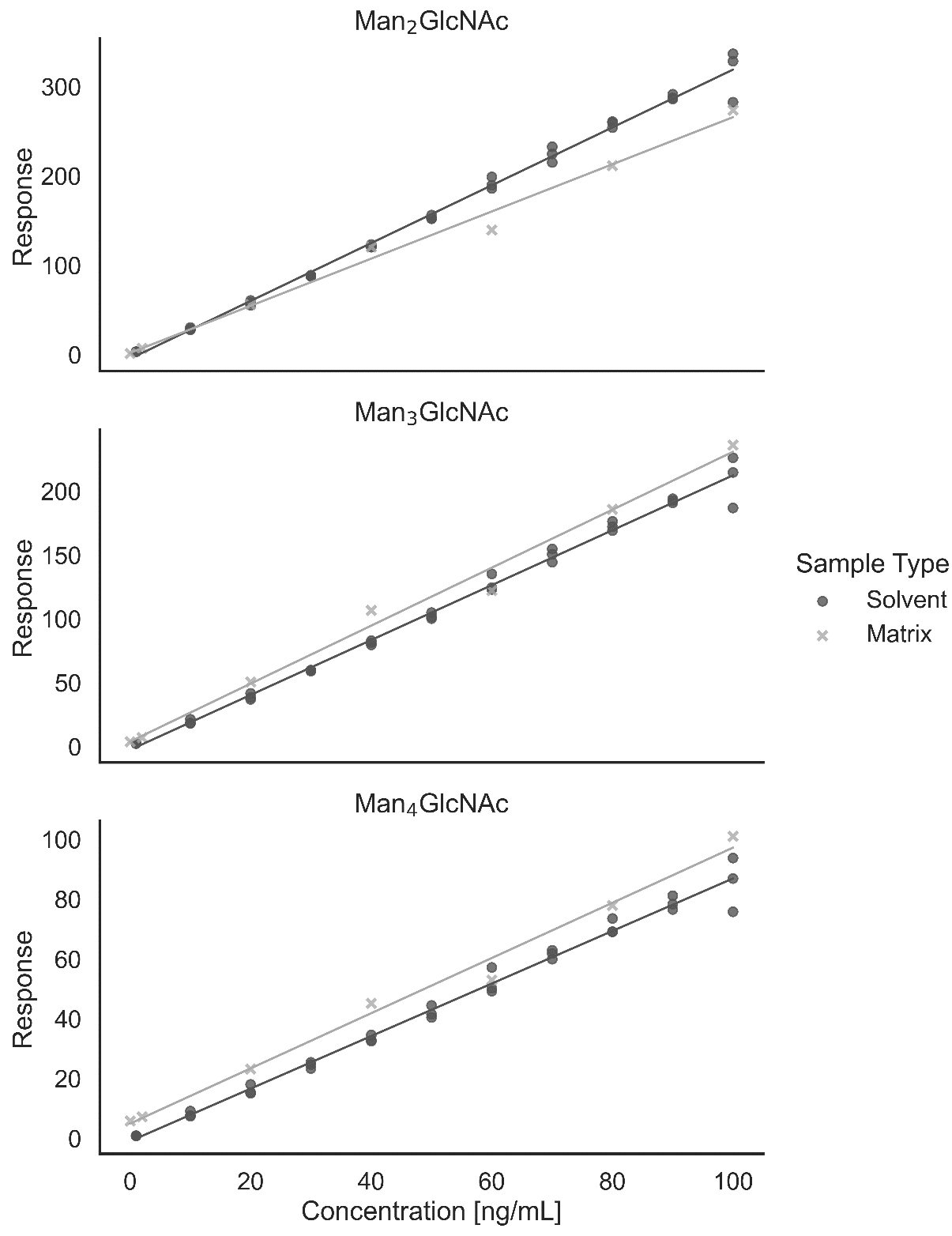

##### **Figure S1:** Linearity assessment for all three biomarkers in solvent and matrix.

##### **Table S5**: Linear regression parameters for Man_2-4_GlcNAc in solvent and matrix.

| Analyte | Solvent Calibration | | Matrix Calibration | |
| --- | --- | --- | --- | --- |
|  | R² | Slope | R² | Slope |
| Man_2_GlcNAc | 0.9973 | 3.15 | 0.9893 | 2.37 |
| Man_3_GlcNAc | 0.9969 | 2.10 | 0.9893 | 2.27 |
| Man_4_GlcNAc | 0.9959 | 0.86 | 0.9894 | 0.92 |

#### Carry-over

We assessed the carry-over by injecting a solvent blank immediately after a calibration standard with a concentration of 100 ng/mL for all three analytes. If a peak was detected, the peak area was divided by the area of the peak at the analyte’s LLOQ. Carry-over had to be < 20 % for the analytes and < 5 % for the internal standard, as set forth by the method validation guideline.

For the IS, no peak was detected, resulting in a carry-over of 0 %. For Man_2_GlcNAc, Man_3_GlcNAc, and Man_4_GlcNAc, a small peak was detected, corresponding to a carry-over of 0.6 % each for Man_2_GlcNAc and Man_3_GlcNAc, and 1.8 % for Man_4_GlcNAc. These values are within the acceptance criteria. Therefore, the carry-over is validated successfully for all four compounds.

#### Limit of detection and quantification

The LOD and LOQ were determined by 10-fold analysis of a sample from an unaffected individual against a calibration curve and calculated as multiples of the resulting standard deviation. Formulas 1 and 2 are used for their calculation:

(1)	$LOD=3.3*\sigma$

(2)	$LOQ=10* \sigma$

where *σ* is the standard deviation of the ten measurements. The resulting LODs and LOQs for each analyte are presented in Table S6. With LODs of 0.25 µmol/L and below, this assay proves highly sensitive for the analysis of all three oligosaccharide biomarkers in DBS.

##### **Table S6**: LOD and LOQ for Man_2-4_GlcNAc in DBS.

| Analyte | LOD  [µmol/L] | LOQ  [µmol/L] |
| --- | --- | --- |
| Man_2_GlcNAc | 0.09 | 0.29 |
| Man_3_GlcNAc | 0.07 | 0.22 |
| Man_4_GlcNAc | 0.25 | 0.76 |

*LOD,* limit of detection; *LOQ,* limit of quantification*.*

#### Accuracy and precision

To determine accuracy and precision, we spiked EDTA-blood of a healthy volunteer with three different concentrations of the three analytes. These concentrations represented a low spike level at 1 µg/mL blood, a medium spike level at 5 µg/mL blood, and a high spike level at 12.5 µg/mL blood. These concentrations correspond to 1.8 µmol/L for Man_2_GlcNAc, 1.4 µmol/L for Man_3_GlcNAc, and 1.2 µmol/L for Man_4_GlcNAc at the low level. At the medium level, concentrations are 9.2 µmol/L, 7.1 µmol/L, and 5.7 µmol/L; and at the high spike level, the concentrations are 22.9 µmol/L, 17.7 µmol/L, and 14.4 µmol/L for Man_2_GlcNAc, Man_3_GlcNAc, and Man_4_GlcNAc, respectively. These concentrations correspond to 3.2 ng/punch (*low*), 16 ng/punch (*medium*), and 40 ng/punch (*high*), respectively for all analytes, based on the assumption that a DBS punch with a diameter of 3.2 mm contains 3.2 µL of blood. We used these samples to perform intra-assay (*n* = 6) and inter-assay (*n* = 6) measurements. The inter-assay measurements were conducted on three different days by two different scientists. Accuracies are calculated by Formula 3:

(3)	$Accuracy= \frac{c_{sample}-c_{blank}}{c_{spiked}}$,

where c_sample_ is the measured concentration of the spiked samples, c_blank_ is the measured concentration of the non-spiked sample, and c_spiked_ is the nominal concentration that was added to the spiked samples. Precision was calculated as the relative standard deviation of the intra-assay accuracies. According to the method validation guidelines, acceptance criteria are ± 15 % of the nominal concentration, or 85 – 115 %, for accuracies and precision below 15 % relative standard deviation for each spike level. The results are displayed in Table S7.

##### **Table S7**: Results for the accuracies and precisions of Man_2-4_GlcNAc.

| Analyte | Accuracy  [%] | | | Intra-assay precision  [% RSD] | | | Inter-assay precision  [% RSD] | | |
| --- | --- | --- | --- | --- | --- | --- | --- | --- | --- |
|  | L | M | H | L | M | H | L | M | H |
| Man_2_GlcNAc | 80 | 94 | 96 | 10 | 4 | 7 | 14 | 7 | 6 |
| Man_3_GlcNAc | 103 | 122 | 125 | 9 | 4 | 7 | 20 | 13 | 10 |
| Man_4_GlcNAc | 114 | 122 | 128 | 17 | 5 | 9 | 31 | 13 | 8 |

*RSD,* relative standard deviation;

*L*, lower spike level; *M*, medium spike level; *H*, high spike level*.*

For Man_2_GlcNAc, the accuracy of the low spike level (80 %) does not meet the acceptance criteria, while the other two accuracies along with all precisions are within the acceptance criteria of the method validation guideline. At the low spike level, the mild deviation from the acceptance criteria is accepted, as the impact of the slightly diminished recovery is negligible in comparison to the very clear separation of Man_2_GlcNAc concentrations between the control cohort and the cohort of untreated AM patients, which should ensure that there are no adverse effects on the diagnostic power of this assay. Similarly, treatment monitoring is unlikely to be significantly affected as it seems highly improbable that a 5 % deviation in biomarker concentration would lead to a treatment adjustment.

Regarding quantification of Man_3_GlcNAc and Man_4_GlcNAc, multiple accuracies and precisions do not fulfill the acceptance criteria. This can be explained by the semiquantitative nature of the method, which is based on a chemically distinct, isotope-labeled standard (Man_2_GlcNAc-IS) with different elution properties. Overall, their analysis by this method did not show satisfactory accuracy, and they have been omitted from further validation and interpretation.

As a result, the validation of this method demonstrates satisfactory accuracy and precision for Man_2_GlcNAc.

#### Stability

To assess the stability in matrix, we stored DBS of the low and high spike levels (*see* Accuracy and precision) at room temperature (20 °C) and in the freezer (-20 °C) and analyzed their Man_2_GlcNAc concentrations weekly over a period of eight weeks and once every four to eight weeks, subsequently. As per the method validation guideline, stability is satisfactory if the measured concentration does not exceed a span of ±15 % of the nominal concentration. However, contrary to the assumption of the guideline, no blank sample or sample with nominal concentration is available, *i.e.*, there are no human DBS samples without detectable amounts of Man_2_GlcNAc. Hence, we used a spiked sample of a healthy volunteer and determined baseline concentration by our method. To account for the imprecision of the baseline determination, we added the inter-assay imprecision of the respective spike level to the method validation guideline’s acceptance criteria. As a result, the acceptance criteria for Man_2_GlcNAc were 71 – 129 % of the baseline concentration for the low spike level and 79 – 121 % of the baseline concentration for the high spike level. The results of the sample stability assessment are shown in Figure S2.

Up to 33 weeks (231 days), all stability assessments are within the acceptance criteria with the exception of one measurement of the high sample stored at -20 °C (day 176). This measurement was treated as an outlier, since the next and all subsequent measurements were well within the acceptance criteria.

From week 33 (day 231) on, the low concentration sample stored at room temperature resulted in recoveries below the acceptance criteria (< 71 %). This finding, as well as the initial decrease of approximately 20 %, may be attributable to residual activity of alpha-mannosidase in the blood of the healthy volunteer, which has less relative impact on the higher Man_2_GlcNAc concentrations in a high concentration sample.

Based on the assumption that sample turnaround is faster than 33 weeks in routine laboratory environments for both diagnosis and treatment monitoring, we do not consider the effects of a decline of the biomarker concentration beyond 33 weeks of storage to be of practical relevance. This might, at most, have played a role in this study in samples from untreated AM patients that had been stored for a longer period; however, we can assume that these samples do not exhibit sufficient alpha-mannosidase activity to cause such a storage effect.

In conclusion, the long-term sample stability of Man_2_GlcNAc has been demonstrated for up to 41 weeks of storage at -20 °C. Furthermore, our results demonstrate that prolonged shipment of samples is possible without the need for refrigeration and without affecting analysis results. Thus, the analyte stability for this method is validated successfully.

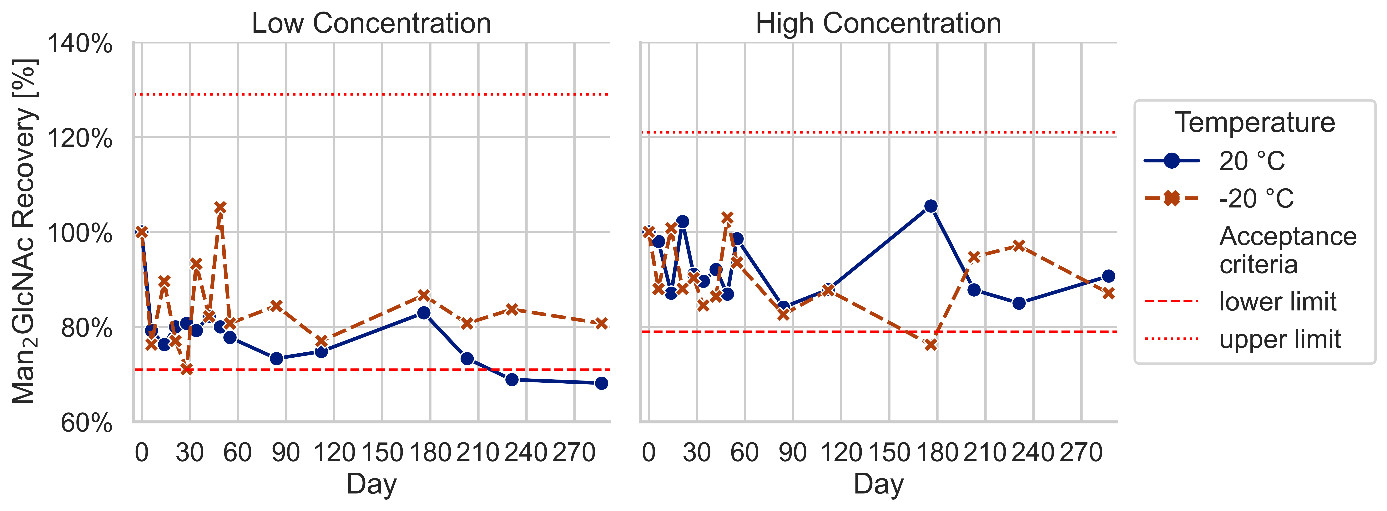

**Figure S2:** Long-term stability of Man_2_GlcNAc in DBS.

#### Matrix effect

According to the method validation guideline, the matrix effect is the alteration of the analyte response due to interfering components in the sample matrix. To assess this effect, we analyzed six randomly selected residual control samples without addition of Man_2_GlcNAc (*blank*), and after addition of two different amounts of Man_2_GlcNAc after extraction. The two different concentrations were comprised of a low concentration corresponding to 2 ng/mL in the sample extract, or 0.57 µmol/L in blood (*low*), and a high concentration corresponding to 20 ng/mL in the sample extract, or 5.7 µmol/L in blood (*high*). The three different kinds of samples (*i.e.*, blank, low, and high) of all six individuals, were analyzed in triplicate. Thereafter, we calculated the accuracy and precision in accordance with the calculation described previously (*see* Accuracy and precision). Acceptance criteria were accuracies within 85 – 115 % and precisions, expressed as relative standard deviation, below 15 %. The results of the matrix effect assessment are provided in Table S8.

**Table S8:** Matrix effect for Man_2_GlcNAc in samples from six individuals matrices for low and high spiking levels.

| Sample | Low concentration (n = 3)  (mean ± RSD)  [%] | High concentration (n = 3)  (mean ± RSD)  [%] |
| --- | --- | --- |
| 1 | 102 ± 4 | 88 ± 3 |
| 2 | 107 ± 4 | 90 ± 2 |
| 3 | 107 ± 2 | 90 ± 1 |
| 4 | 105 ± 2 | 91 ± 2 |
| 5 | 110 ± 3 | 86 ± 5 |
| 6 | 110 ± 2 | 87 ± 5 |

*RSD*, relative standard deviation.

For all six individual samples, the calculated accuracies and precisions are within the acceptance criteria of 85 – 115 % and below 15 % relative standard deviation, respectively. In conclusion, the matrix effect of our method is negligible and has been validated successfully.

#### Analytical specificity

The method validation guideline defines the analytical specificity of a method as the ability to detect the analyte and differentiate it from other substances. In general, liquid chromatography (LC) tandem-mass spectrometry (MS/MS) assays exhibit a high level of analytical specificity due to the combination of analyte separation by LC with MS/MS detection, where the combination of precursor and product ions is often specific for the compound tested. Previous work of our group on a method for oligosaccharide analysis in urine has shown that method specificity is compromised by other isobaric oligosaccharides present in urine [3]. Therefore, it is very likely that these oligosaccharides, if present, would also affect specificity in DBS. However, like in urine, interfering oligosaccharides would only impact biomarker analysis in DBS if present in high concentrations. In urine, this resulted in highly elevated quantifier-qualifier-ratios (Q/q > 20, in comparison to a Q/q of 1.2 for a standard solution of Man_2_GlcNAc). In DBS, no sample exhibited elevated amounts of Man_2_GlcNAc (*i.e.*, > 1 µmol/L) in combination with an elevated Q/q. Overall, no adverse effect on the analytical specificity was observed during method development and validation. In conclusion, the analytical specificity of this method has been validated successfully.
